# Mathematical modeling of COVID-19 containment strategies with considerations for limited medical resources

**DOI:** 10.1101/2020.04.17.20068585

**Authors:** Brydon Eastman, Cameron Meaney, Michelle Przedborski, Mohammad Kohandel

**Affiliations:** Department of Applied Mathematics, University of Waterloo, ON N2L 3G1

## Abstract

The outbreak of SARS-CoV-2 in China has spread around the world, infecting millions and causing governments to implement strict policies to counteract the spread of the disease. One of the most effective strategies in reducing the severity of the pandemic is social distancing, where members of the population systematically reduce their interactions with others to limit the transmission rate of the virus. However, the implementation of social distancing can be difficult and costly, making it imperative that both policy makers and the citizenry understand the potential benefits if done correctly and the risks if not. In this work, a mathematical model is developed to study the effects of social distancing on the spread of the SARS-CoV-2 virus in Canada. The model is based upon a standard epidemiological SEIRD model that has been stratified to directly incorporate the proportion of individuals who are following social distancing protocols. The model parameters characterizing the disease are estimated from current epidemiological data on COVID-19 using machine learning techniques. The results of the model show that social distancing policies in Canada have already saved thousands of lives and that the prolonged adherence to social distancing guidelines could save thousands more. Importantly, our model indicates that social distancing can significantly delay the onset of infection peaks, allowing more time for the production of a vaccine or additional medical resources. Furthermore, our results stress the importance of easing social distancing restrictions gradually, rather than all at once, in order to prevent a second wave of infections. Model results are compared to the current capacity of the Canadian healthcare system by examining the current and future number of ventilators available for use, emphasizing the need for the increased production of additional medical resources.

## 1 Introduction

On January 11, 2020, China reported the first death due to a novel strain of coronavirus, now officially named SARS-CoV-2, to the World Health Organization (WHO) [1]. To date, the origin of the virus remains uncertain; however, it is thought to have originated in a wet market in Wuhan, China via inter-species transmission to humans. By January 20, 2020, the number of infections in China had sharply risen to 278, including six deaths, and several cases had also been confirmed in Japan, South Korea and Thailand [1]. The virus quickly spread to North America through intercontinental travel: the CDC reported the first case in the State of Washington, United States on January 21, 2020 [2], and four days later Canada confirmed its first case in Toronto, Ontario [3].

As of mid-April 2020, close to two million people have been infected globally by SARS-CoV-2 and over 120,000 people have died as a result of complications from COVID-19, the disease caused by the virus [4]. The virus is now affecting people in 210 countries and territories around the globe. To attenuate the spread of the virus and mitigate the impact on the medical system, several different social distancing strategies have been implemented in countries around the world. These strategies range from quarantining infected individuals and those returning from international travel, to orders to avoid public parks and facilities, to prohibiting gatherings of more than two people (with exceptions for family members). In South Korea, for instance, a recent report has indicated that social distancing protocols were responsible for reducing the effective *R*_0_ value in various cities (below 1 in some cases) by estimating social distancing effects via traffic or metro data [5]. In addition, in some countries, all day cares, educational facilities, and non-essential businesses have been ordered temporarily closed by government officials, and citizens have been ordered to stay home, except to go shopping for essential supplies, such as groceries and prescriptions.

Data obtained by tracking mobile phone locations [6, 7] indicates that a fair proportion of people are abiding by social distancing policies in North America. In particular, data collected for several weeks between March 2020 and mid-April 2020 indicate that mobility toward the workplace has decreased between 24% and 52% in different parts of North America while mobility toward retail and recreation has decreased between 29% and 70% [6, 7]. These data raise the questions of how these policies have modulated the timeline of SARS-CoV-2 outbreaks, how abiding by social distancing policies have mitigated the overload to the medical system, and how sensitive these effects are to the proportion of the population abiding by the policies.

To address these and other questions, we developed a mathematical model to study the effects of social distancing on the spread of SARS-CoV-2, with an emphasis on the policies implemented in Canada as a focused example. First, using publicly-available data from several online repositories along with machine learning techniques, we fit the temporal evolution of infection counts, total deaths, and total recoveries in 80 different countries to an SEIRD infection model. These fits provided insight into the biologically and socially-relevant parameter ranges for the model. Next, we sampled these parameter ranges to simulate the dynamics of SARS-CoV-2 spread in provinces within Canada using the Distancing-SEIRD model developed in this work. Finally, we studied the disease outlook and the burden on the medical system for different social distancing strategies.

The manuscript is organized as follows. In Section 2, we describe the models used in this work and give a brief discussion of the techniques used to estimate the SEIRD model parameters to fit the temporal SARS-CoV-2 data from several countries. In Section 3, we present our results and give a discussion of the significant model predictions. Finally, in Section 4 we provide some concluding remarks and future research directions. The details of mathematical model and parameter estimations are provided in the Supplementary Information.

## 2 Methods

We consider two models of infectious disease, the full technical details of which can be found in the Supplementary Information, Section A. The first model, the SEIRD model, is illustrated in Figure 1. In this model, we assume that all people fall into one of five categories: susceptible, exposed, infected, recovered or deceased. The “susceptible” category refers to people who have never had the virus (and therefore lack antibodies to defend against it). The “exposed” individuals are those who have caught the virus from an infected individual but who have not yet become contagious themselves. The “infected” individuals are people who have the virus and are now contagious. The “recovered” individuals are people who have gone through the whole life-cycle of the virus and survived. We assume that recovered individuals now have a prepared immune response and will never catch the virus again. Finally, the “deceased” class represents the cumulative number of fatalities as a result of COVID-19 disease. Note that some individuals may never leave the susceptible compartment during an entire pandemic outbreak (representing people who do not contract the virus during an outbreak). However, once someone enters the exposed compartment, they will irreversibly progress through the infected compartment before ending as either recovered or deceased.

**Figure 1:**
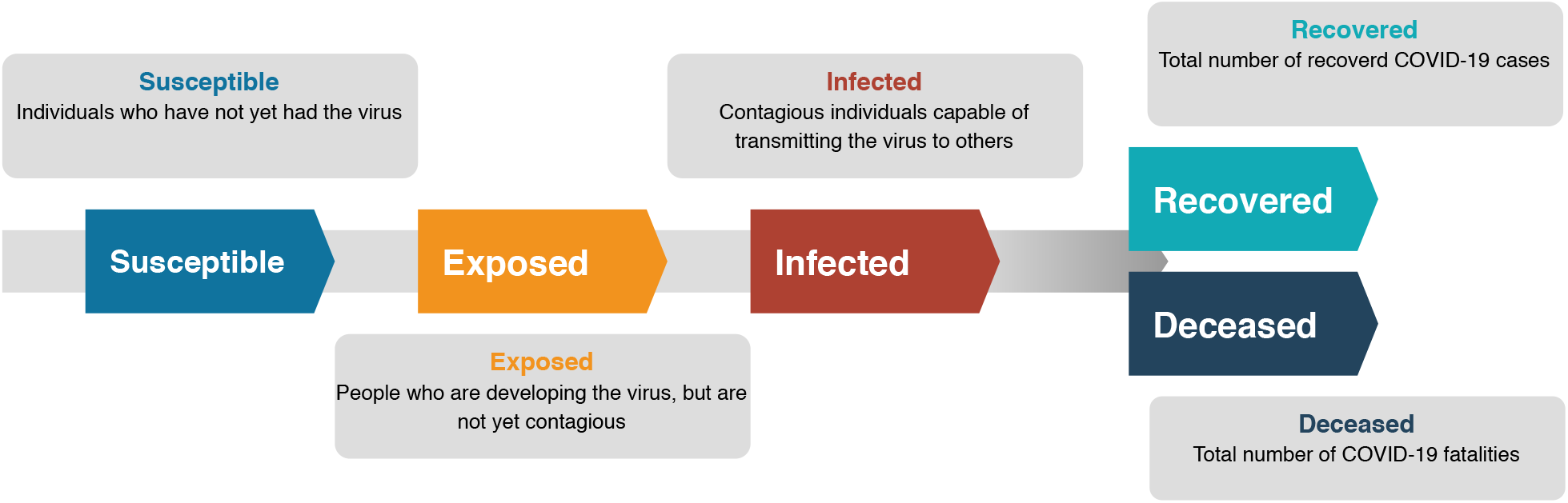
Schematic of the standard SEIRD epidemiological model. The population is divided into five compartments: susceptible, exposed, infected, recovered, and deceased.

The second model we consider is a proposed model of social distancing combined with SEIRD dynamics (details in the Supplementary Information, Section A) as illustrated in Figure 2. This model is an extension of the SEIRD model described above which we refer to as the Distancing-SEIRD model. To incorporate social distancing, we further split the susceptible, exposed, and infected compartments into two types: distancers and mixers. Distancers represent people who are following social distancing protocols. As a result, susceptible distancers have less contact with infectious individuals and are less likely to become sick themselves. Furthermore, infectious distancers are less likely to pass the virus on to someone else as a result of their lower contact rate. Conversely, mixers are more likely to contract and spread the infection as a result of their increased motility.

**Figure 2:**
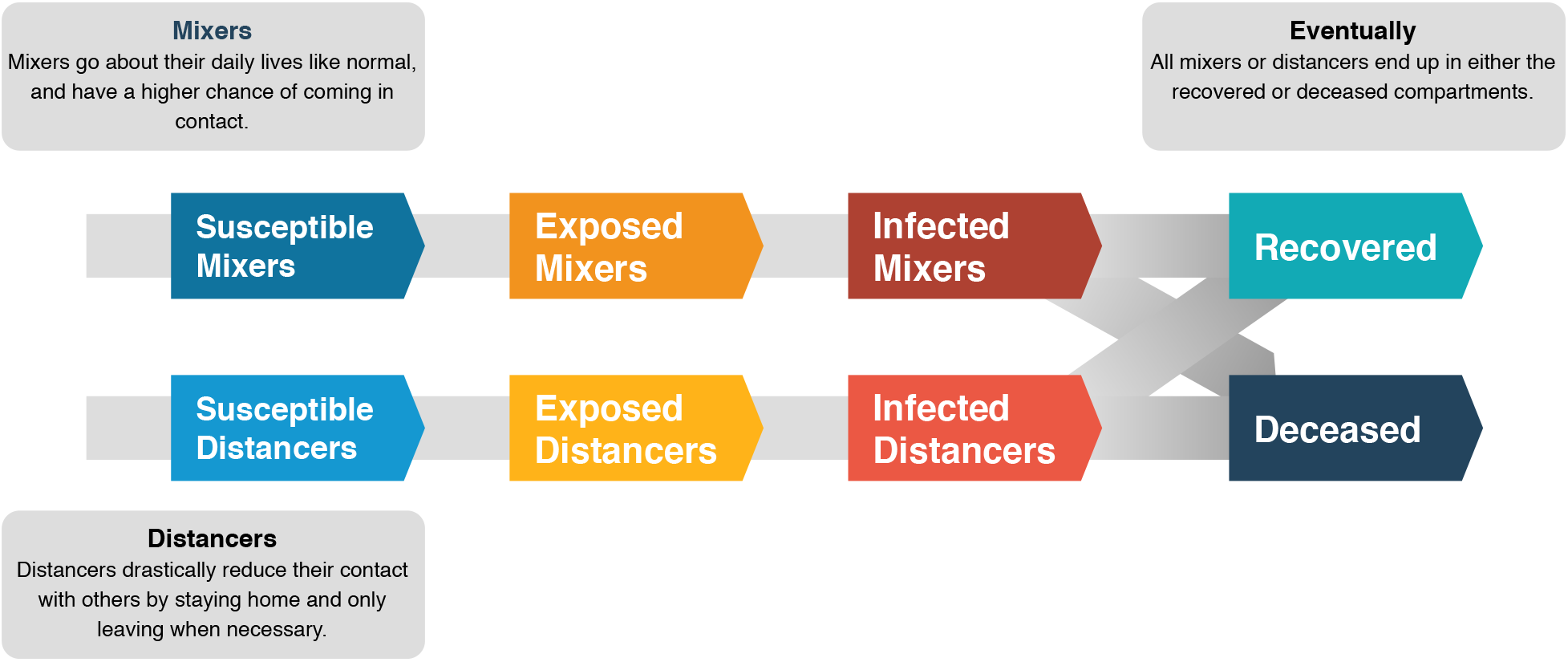
Schematic of the proposed Distancing-SEIRD model. The model is similar to the standard SEIRD model except the susceptible, exposed, and infected compartments are further stratified into two compartments each based on social distancing behavior.

Both of these models require approximating the value of various mathematical parameters. This parameter fitting process is described in detail in the Supplementary Information, Section B.2. The process hinges fundamentally on finding reasonable bounds for the parameters as informed by the epidemiological data. The data we use is collated by the Johns Hopkins University Center for Systems Science and Engineering [8]. With those parameters discovered we can use the models to forecast the spread of SARS-CoV-2 in various populations. In particular, by construction of the Distancing-SEIRD model (as discussed in Figure 2), we are able to investigate the efficacy of social distancing on attenuating the strain on the healthcare system due to COVID-19.

As powerful as these prediction methods are, all modelling is subject to assumptions. For instance, neither model considers how the burden on the healthcare system directly affects the case fatality rate. In practice, we expect that if the healthcare system were to become overburdened as a result of a large number of simultaneous infections, the case fatality rate would increase. This would happen as the result of health care practitioners being unable to provide care for all serious patients due to the sheer volume of cases. As a result, even with effective triage procedures, we would anticipate more fatalities from COVID-19 in an overburdened healthcare system than in one with extra resources, even when the total case count is constant over a period. Further, in the two models presented we ignore the role of random events, assume that the population is well mixed, and assume that the spread of the virus in each geographical region behaves like a single outbreak. In reality, random events can influence the spread of the virus in profound ways, populations are quite heterogeneous and not well mixed, and, especially in the early stages of an outbreak, the spread of the virus in a province, state, or country is often characterized by multiple smaller outbreaks happening out of sync with one another (such as in various cities within a province). As an epidemic outbreak progresses, the validity of these assumptions changes. For instance, in the early days of a viral outbreak in a particular province, we might observe smaller, discrete outbreaks in multiple cities evolving without any influence from one another; however, as those outbreaks grow in size, they eventually overlap and further behave as a single outbreak. It is for these exact reasons that in the Canadian data we only provide results for the two provinces that, at the time of this writing, have the largest outbreaks. Namely, the Canadian provinces of Quebec and Ontario.

## 3 Results and discussion

Here we present the main findings of the study and the predictions of the standard SEIRD and Distancing-SEIRD models. First, we depict the short-term and longer-term fits to the country-wide data for Canada and China, as well as the fits to COVID-19 epidemiological data from Quebec and Ontario. We use the parameter fits to inform the Distancing-SEIRD model to ultimately make long-term predictions about the impact of social distancing strategies on the dynamics of the pandemic in Quebec and Ontario.

The fits to the epidemiological data for China and Canada, obtained with the standard SEIRD model, are depicted in Figure 3. The full fitting process is described in the Supplementary Information Section B.2. The process involves estimating key parameters from the raw epidemiological data and using machine learning techniques to optimize the remaining parameters.

**Figure 3:**
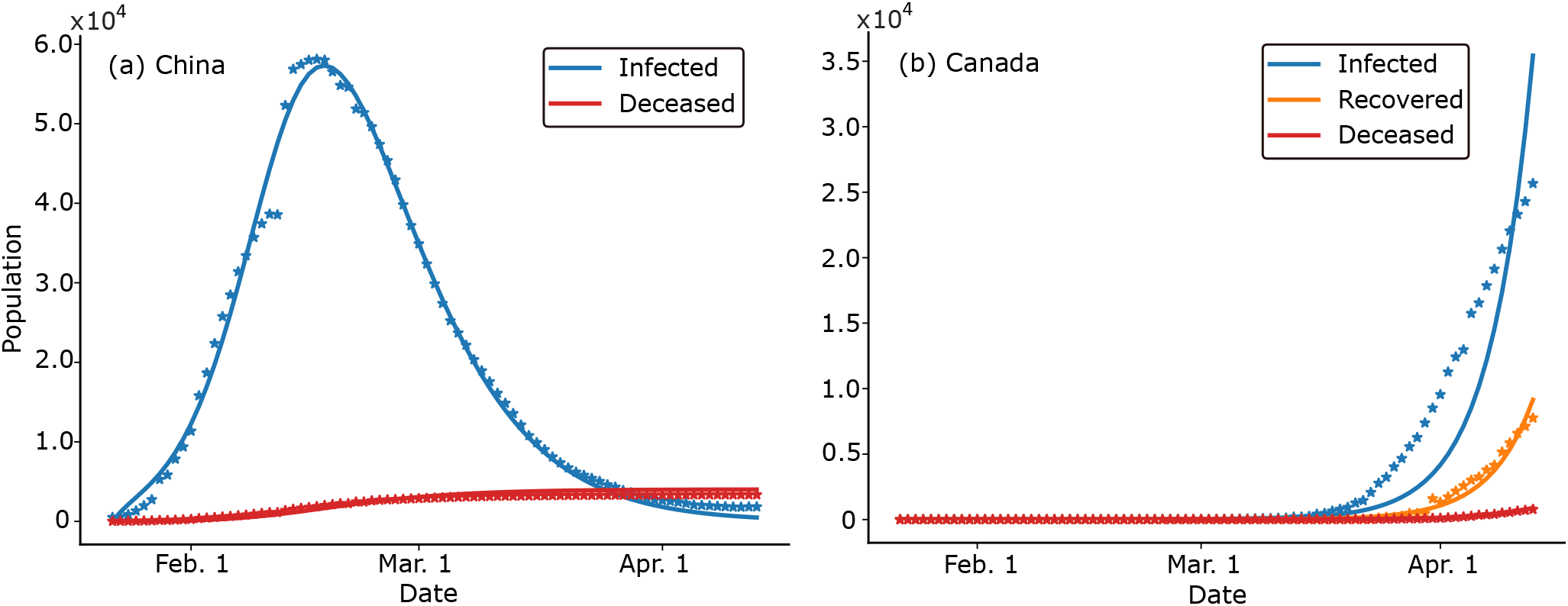
Temporal evolution of currently infected individuals, total death count, and total number of recovered individuals in (a) China and (b) Canada. Reported data are shown as asterisks. Predictions of the mathematical model, Eq. (A.1), are shown as solid lines. Infected individuals are marked in blue, recovered in orange, and deceased in red.

Figure 3 illustrates that the SEIRD model captured the overall trends in the number of infected individuals, the total number of deaths, and the number of recovered individuals. Importantly, with a suitable and realistic choice of parameters, the model captured the peak and the tail of the longer-term temporal data from China. Note that while the data fits for the infected curve for Canada (Figure 3 b) show quantitative disagreement, the qualitative dynamics of the curve are captured. Importantly, the deceased and recovered curves fit the data quite well. As discussed in Section 2, in the early dynamics of the epidemic at the national or provincial level the epidemic does not evolve as a single outbreak, but rather as the sum of multiple outbreaks in multiple cities with temporal delay. As a result, a model like Eq. (A.1) ought not to perfectly fit these early dynamics. In fact, if we did not set some of the parameters with epidemiological insights (as described in the Supplementary Information Section B.2) we could achieve stricter quantitative fits of the early dynamics of the epidemic at the cost of long term prediction. Parameter values for the Canadian provinces of Quebec and Ontario are provided in Table 1.

**Table 1:** Parameter Values for the SEIRD Model Eq. (A.1) for various locales.

| | $\beta$ (person<sup>-1</sup> day<sup>-1</sup>) | $\gamma$ (day<sup>-1</sup>) | $\alpha$ (day<sup>-1</sup>) | $\delta$ (day<sup>-1</sup>) |
| --- | --- | --- | --- | --- |
| China | $3.501 \times 10^{-6}$ | $1.257 \times 10^{-1}$ | $1.467 \times 10^{-1}$ | $2.404 \times 10^{-3}$ |
| Canada | $1.337 \times 10^{-8}$ | $1.747 \times 10^{-1}$ | $6.424 \times 10^{-2}$ | $6.327 \times 10^{-3}$ |
| Ontario | $7.017 \times 10^{-8}$ | $1.002 \times 10^{-1}$ | $1.217 \times 10^{-1}$ | $1.199 \times 10^{-2}$ |
| Quebec | $1.880 \times 10^{-6}$ | $1.000 \times 10^{-1}$ | $9.194 \times 10^{-2}$ | $9.051 \times 10^{-3}$ |

Using the best fit parameters obtained from the procedure outlined in the Supplementary Information B.2, we sought to predict the effectiveness of several social distancing strategies on mitigating the severity of the pandemic. To this end, we expanded the SEIRD model to directly capture the effects of a proportion of the population adhering to the social distancing guidelines, see Eq. (A.2). Mathematically, this involved first stratifying the susceptible, exposed, and infected compartments of the SEIRD model, Eq. (A.1), based on social distancing behaviour. The resulting system of coupled ODEs was solved numerically using Euler’s method with a time step of 0.1 days, to obtain the temporal evolution of each population compartment over the relevant time period.

While cell phone tracking data has been used to measured the approximate proportion of the population in Canada that is adhering to social distancing in different provinces [6], it is not clear precisely how these numbers translate into a reduction in disease transmission rates. Thus, this work required assumptions to be made about the effects of social distancing on the rate of disease transmission in the Distancing-SEIRD model. Specifically, it was assumed throughout the work that the disease transmission rate between two individuals who are not social distancing, i.e. between two mixers, is 20 times higher than between a mixer and a distancer, and 100 times higher than between two distancers. For specific parameter values, see Table 1 in the Supplementary Information B.2.

For numerical simulations, it was also necessary to make an assumption about the effective day on which social distancing began in each province. It is likely that some degree of social distancing began prior to the widespread adoption in the population and the enforcement of social distancing policies by government officials as the public became progressively more aware of the severity of the pandemic. For consistency and simplicity, in the Distancing-SEIRD model, the adherence to social distancing among the population was assumed to have begun on the date of provincial declaration of a state of emergency - March 17, 2020 for Ontario and March 27, 2020 for Quebec. Under these assumptions, the Distancing-SEIRD model was solved for Ontario and Quebec, where day one of the simulations was chosen to be the date of the first confirmed SARS-CoV-2 case in Canada, which is January 25, 2020 [4]. Thus, social distancing was assumed to start on day 62 of the pandemic in Quebec, and day 52 of the pandemic in Ontario.

The proportion of the population practicing social distancing was found to be a crucial indicator for how effective the social distancing strategy is. To illustrate this point, the Distancing-SEIRD model was used to predict the total number of expected deaths for different levels of social distancing in Quebec and Ontario. The results are depicted in Figure 4 for Quebec and Ontario, where the period of social distancing was taken to last six months. In this figure, the total number of deaths immediately after social distancing is stopped, as well as two years after social distancing first started, are shown.

**Figure 4:**
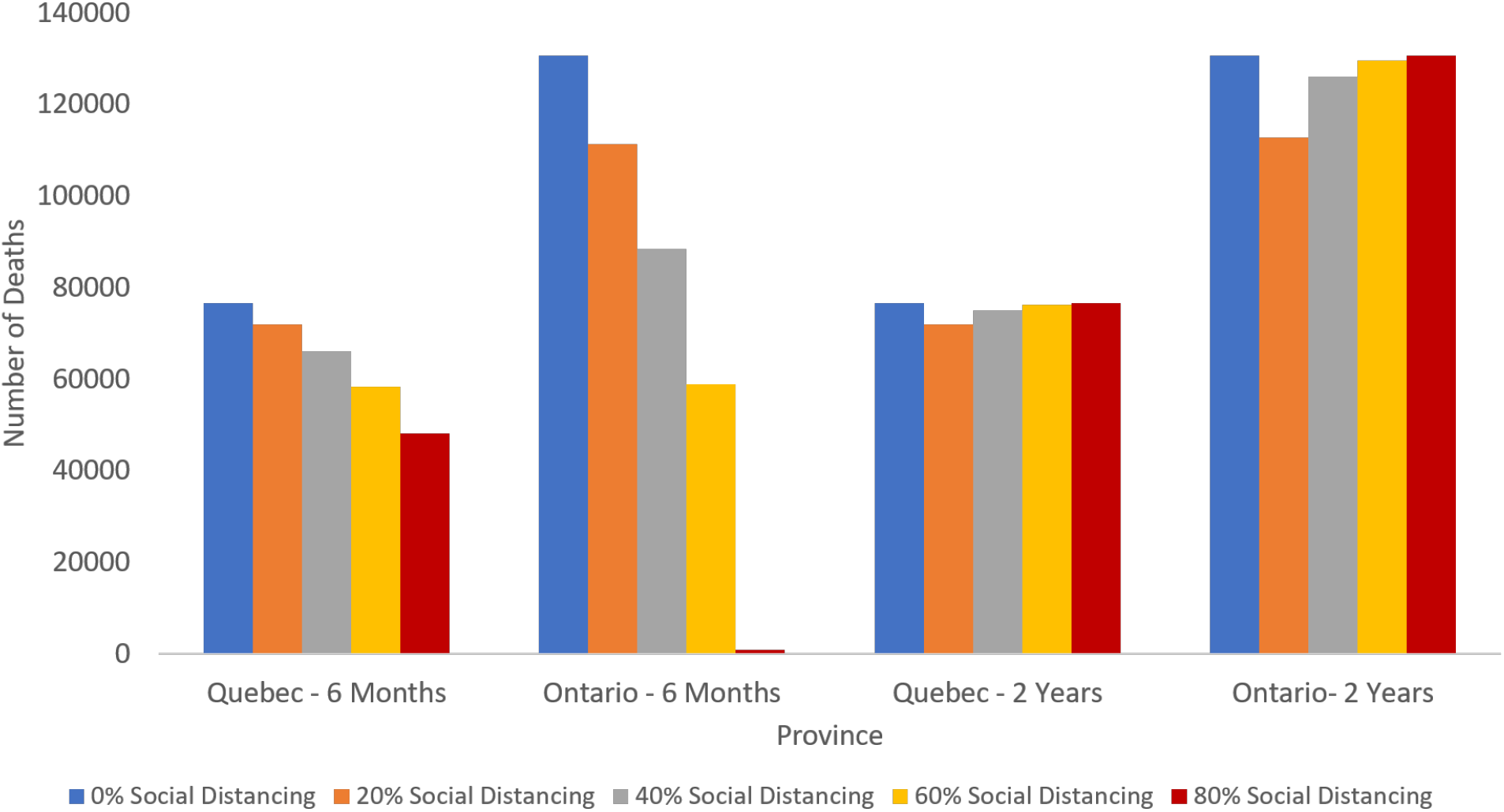
Estimated total number of deaths in Quebec and Ontario due to COVID-19, six months and two years of social distancing for various levels of social distancing adherence. The results indicate that after two years, the total death count is approximately equal for all social distancing behaviours. However, the total deaths after six months decrease drastically with increased social distancing, suggesting a flattening of the infection curve. Such a flattening implies a gradual accumulation of deaths, which better enables the health care system to deal with the wave of infections.

Specifically, we see from Figure 4 that in Ontario, the Distancing-SEIRD model predicted the total number on deaths over a six month period with no social distancing to be as high as 130,000 people. In contrast, the most strict case of social distancing, with 80% adherence, led to a drastic reduction in the death toll - to under 1000 total deaths over the first six months. Unfortunately, Quebec has experienced higher numbers of COVID-19 cases, indicating that the progression of the pandemic is further along than in Ontario. Consistently, the Distancing-SEIRD model predicted that social distancing measures will lead to a less drastic reduction in deaths compared to Ontario. Despite this reduction, the importance of social distancing measures is still evident in Quebec: the simulations indicated that strict social distancing measures could lead to nearly 30,000 less deaths over a six-month span, compared to no social distancing. Studies attempting to estimate the level of adherence to social distancing policies have reported that approximately 60% of people are practicing social distancing in Ontario and 70% in Quebec [6]. Our Distancing-SEIRD model indicates that if social distancing continues at these levels for a six month period (until mid-September) the combined amount of lives saved in Ontario and Quebec alone is approximately 90,000.

Interestingly, the total number of deaths at two years after the start of social distancing, i.e. at 18 months after social distancing is stopped, was found to be less sensitive to the social distancing policy, as indicated by Figure 4. In other words, the total number of deaths over the two year period was predicted to be approximately the same, regardless of the social distancing policy. However, by flattening the infection curve, social distancing leads to a gradual accumulation in the death toll, reducing the strain on the heath care system. Importantly, inspection of Figure 4 reveals that at the two-year mark, strict social distancing policies with 80% adherence were predicted to lead to a death toll that is most similar to the case of no social distancing. This indicates that the principal effect of a strict social distancing policy is to shift the infection peak. Delaying the onset of the infection peak allows the health care system to prepare for the wave of patients, allowing for the creation of additional supplies so that more patients can receive the necessary care. This stresses the idea that a gradual reduction is social distancing, rather than an abrupt end to the policy, would be most effective at reducing the total death toll.

Another important consideration when devising a social distancing strategy is to carefully select an appropriate time to end social distancing. Obviously, various factors influence such a decision as many people face significant economic, social, and personal hurdles as social distancing is prolonged. The risk of ending social distancing too early is that the pandemic may reach a second peak, again overwhelming the healthcare system and leading to unnecessary deaths. A relevant problem therefore, is to determine the earliest time such that social distancing can be responsibly ended. One way to assess control of the virus is to limit the maximum number of concurrent infections to within the capacity of what the healthcare system can handle. A key limited medical resources in treatment of COVID-19 is mechanical ventilators and unfortunately, Canada, as well as most countries, are unlikely to have enough ventilators in order to meet the demand that the pandemic peak will necessitate. Currently, Ontario and Quebec have an estimated 1810 and 2970 ventilators, respectively [9]. In China, an estimated 20% of COVID-19 patients were hospitalized with 6.1% experiencing severe respiratory failure [10]. This can be used in the SEIRD model for the approximate proportion of COVID-19 patients who require the use of a mechanical ventilator, yielding an estimate of the anticipated ventilator demand at the peak of the pandemic by scaling the predicted infection curve.

In Figure 5, the Distancing-SEIRD model is solved for different social distancing strategies and used to obtain the anticipated ventilator need over time for Quebec and Ontario. Cases simulated include social distancing for 3 months and 6 months with adherence levels of 20%, 40%, 60%, and 80%. In Quebec, increasing the proportion of social distancers reduces the size of the initial peak, shifting the demand to a secondary peak which occurs after social distancing has ended. Naturally, the onset of this resurgence is delayed the longer that social is in effect. In Ontario, a 20% social distancing adherence reduces the size and slightly delays the onset of the initial wave of ventilator demand. For 40% social distancing, the initial wave is further reduced and delayed, but the effect of choosing the proper stop time is clear. Ending social distancing after three months creates a second wave approximately one month later, whereas extending social distancing for six months delays the second wave until the winter, allowing time for the production of additional ventilators in time to meet the demand. For high proportions of social distancing, the main effect becomes delaying the curve, rather than flattening it. Indeed, for 80% social distancing, the ventilator demand curve is nearly identical to the case with no social distancing except for a time-shift until after social distancing has ended. This again stresses the importance of a gradual reduction in social distancing so as to not shock the system.

**Figure 5:**
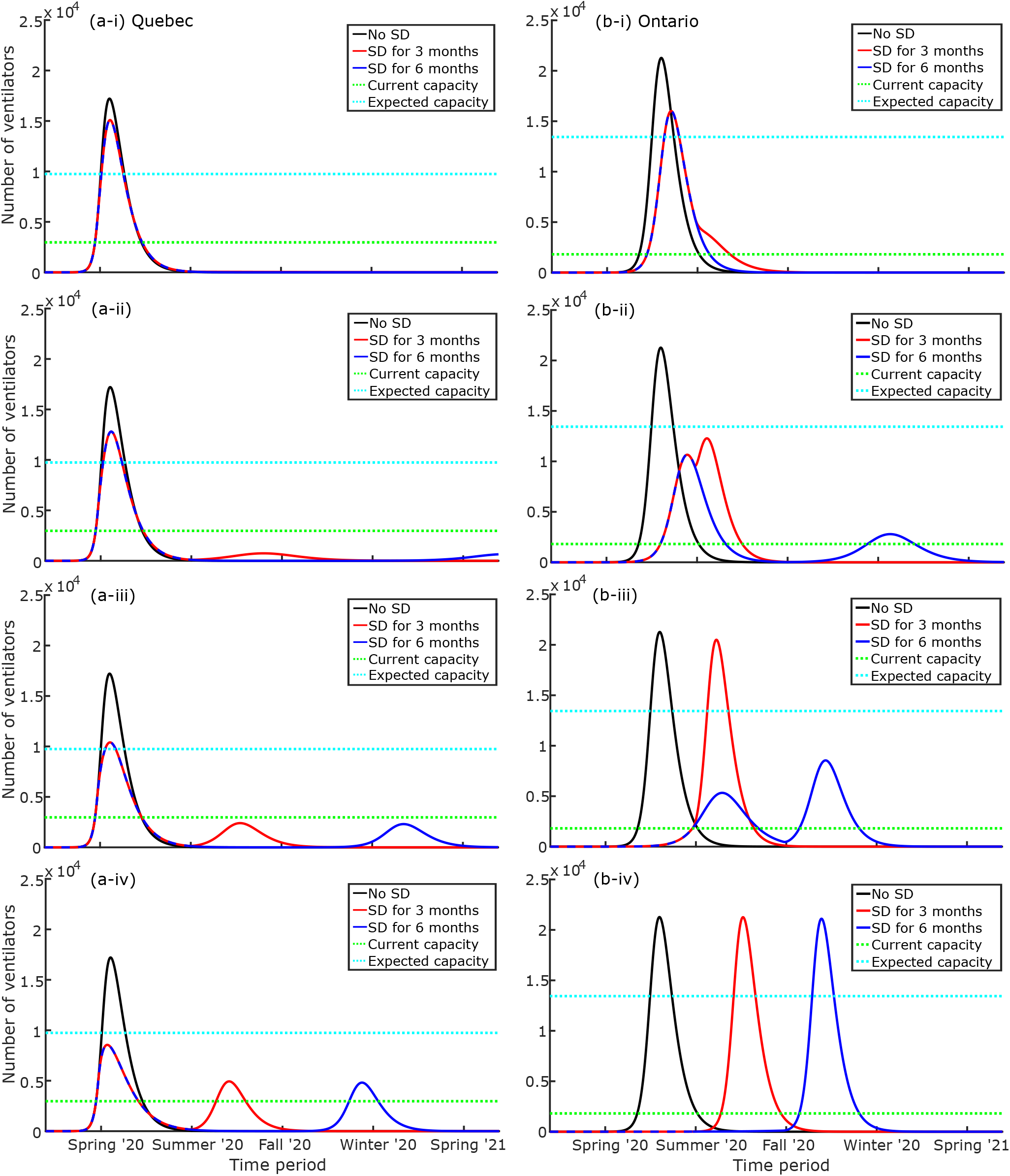
Projected ventilator requirements due to COVID-19 infections in (a) Quebec and (b) Ontario, as a function of time. Infected patients Each plot shows the predictions for no social distancing (SD), 3 months of social distancing, and 6 months of social distancing. Season tick marks represent the start of the respective season. Calculations of expected ventilator capacity are described in the text. The proportion of people adhering to social distancing policies increases from top to bottom: (i) 20%, (ii) 40%, (iii) 60%, and (iv) 80% of the population.

These results are further supported by the behaviour of peak infections vs social distancing time for various policies, as can be seen in Figure 6. In both Quebec and Ontario, the peak number of simultaneous infections decreases as the proportion of social distancers increases. In Quebec, after approximately one month of social distancing, the peak infections reaches a constant value and further social distancing simply delays the onset of the second peak, rather than reducing its size (as can be seen in Figure 5. This is largely because Quebec is further into the pandemic than Ontario, so the size of its first peak cannot be reduced to smaller than its subsequent peaks. In Ontario, the social distancing stop time has a significant impact on the maximum peak infection number. Specifically, for low proportions of social distancing (20% and 40%), the behaviour of the maximum peak behaves much like Quebec; however, for high proportions of social distancing (60% and 80%), the behaviour is qualitatively much different. For 60%, the maximum peak decreases as the length of social distancing increases until it levels-off at approximately six months. For 80%, the maximum peak slowly decreases until levelling-off after approximately 15 months. This is because at 80% social distancing, the main effect is delaying the peak, rather than shrinking it. These results show that choosing the proper time to end social distancing can be crucial in preventing the healthcare system from becoming overburdened.

**Figure 6:**
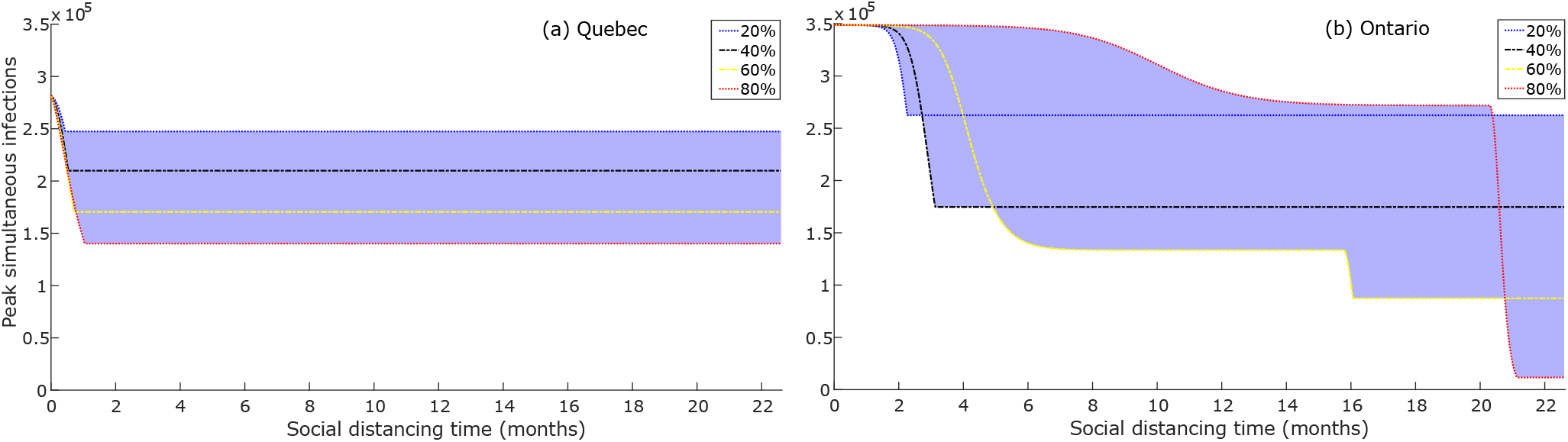
Predicted peak number of simultaneous COVID-19 infections in (a) Quebec and (b) Ontario, as a function of social distancing time. Social distancing time is measured with respect to the start of the implementation of social distancing policies in each province: for Quebec, this was March 27, 2020; for Ontario, this was March 17, 2020. Results are shown for different proportions of individuals adhering to social distancing policies and indicate a qualitative change in outcome for Ontario as the social distancing proportion exceeds approximately 40% of people.

The unfortunate reality is that in no cases were social distancing policies sufficient to keep the ventilator demand of the first wave below the current capacity of the respective provincial healthcare systems. With proper social distancing policies, it is possible to restrict the size of the second peak to below the current capacity, but these cases involve a potentially optimistic vision of the levels of public adherence to social distancing guidelines for a long period of time. Fortunately, the increased need for ventilators has been recognized by government officials and the Canadian government has announced the production of 30,000 new ventilators to combat the pandemic. Assuming these ventilators are divided among the provinces proportionally based on population, we can assess whether this number is sufficient to handle the anticipated demand. Looking at Figure 5, the increased ventilator capacity significantly increases the likelihood that a social distancing strategy will be able remain beneath the capabilities of the health care system.

## 4 Conclusions

Mathematical models of pandemic progression can serve as a useful tool for policy-makers crafting a societal pandemic response. If used correctly, they can also serve as tools for persuading the general public on best practices for controlling the virus’ spread. One of the main tactics the public can employ to combat the virus is social distancing: limiting the amount of social interaction in order to lower the effective transmission rate. Unfortunately, the widespread use of social distancing can cause many economic, social, and personal challenges, leading some to question the value of social distancing in achieving pandemic control. Naturally, questions arise regarding how many people must social distance, and for how long, in order for these tactics to be effective.

In this work, an epidemiological mathematical model, the Distancing-SEIRD model, is developed for use in predicting the effectiveness of various social distancing strategies in Canada. Disease parameters are fitted to the model using machine learning techniques for the specific cases of Ontario and Quebec and used to project the number of deaths and ventilator demand for proposed social distancing strategies. Social distancing strategies are defined by the proportion of the population social distancing and the length of time over which social distancing occurs. Allowing for a proportion of the population to social distance, rather than all-or-nothing, allows for the more realistic case of imperfect social distancing to be modelled.

Our model results show that the social distancing policies already implemented in Ontario and Quebec have saved tens of thousands of lives, and can continue to do so if social distancing remains in place for a large proportion of the population and a sufficiently long time. The main objective in social distancing is to flatten the infection curve, preventing the healthcare system from becoming over-burdened, and minimizing the need for triage. The flatter the pandemic infection curve, the lesser the need for triage, and therefore the more lives that will be saved. Furthermore, social distancing acts not only to reduce the peak infections, but also to delay it - buying time for the increased production of medical resources, testing of pharmaceutical therapies, and the development of a vaccine. Our results also stress the importance of gradually relieving social distancing rather than all at once. A significant decrease in social distancing at once can result in a second wave of infections which can overwhelm the healthcare system.

We hope that the predictions of our model can assist public health officials in crafting policies to lessen the severity of the pandemic, to inform the public on the potential outcomes of the pandemic, and to serve as an impetus for those who are skeptical of following evidence-based social distancing guidelines. Importantly, we stress that the results presented in this work are estimates that are based upon a simplistic mathematical model which relies on imperfect and incomplete data. A more comprehensive mathematical analysis of the pandemic would require a significantly more complex model and a larger set of data to determine key disease parameters. Therefore, the results should be interpreted qualitatively as an educated estimate of what is possible given different levels of public adherence to social-distancing guidelines.

## Data Availability

Not Applicable.

## Supplementary Information

### A Mathematical Models

We consider two models for the spread of SARS-CoV-2. The first model (Model A.1) is an SEIRD model with five compartments: *S*(*t*) is the population of individuals that are susceptible to the virus, *E*(*t*) is the population of individuals who have been exposed to the virus but are not yet contagious, *I*(*t*) is the population of individuals that are infected by the virus, *R*(*t*) is the population of individuals who have recovered from the disease (and thus have immunity), and *D*(*t*) is the number of fatalities from the disease.

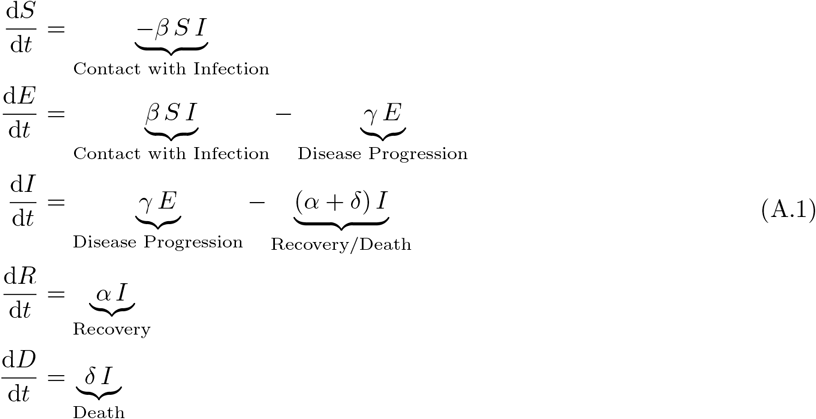

The interpretations of the individual terms in the SEIRD model are given in Eq. (A.1). As illustrated by the set of ordinary differential equations (ODEs) in Eq. (A.1), the SEIRD model depends upon four kinetic parameters. The parameter *β* controls the rate of virus transmission from infected to susceptible individuals. It is determined by the probability of disease transmission as well as the chance of contact, thus it indirectly incorporates the basic reproduction number, *R*_0_, of the virus and the proportion of individuals who are social distancing. The mean latent period, which is the average length of time between exposure to the virus and the point at which an individual becomes contagious, is given by *γ*^−1^. The rate at which infectious individuals are removed from the disease (either via recovery or death) is given by (*α* + *δ*), thus the value of (*α* + *δ*)^−1^ is the mean length of time an infected individual is contagious before they either recover or die from the disease. The five compartments are subjected to the constraint *S* + *E* + *I* + *R* + *D* = *N*, where *N* is the total population. We point out that the state equations are considered to represent direct counts of the population size in this work, rather than proportions of the population. Note that, due to the form of the equations, the parameter *β* in Model 1 includes division by the total population size *N*.

Here we propose a Distancing-SEIRD model, referred to as Model 2, that is similar to the first SEIRD model, except that it now directly takes into account the proportion of individuals who are social distancing. In Model A.2, the *S, E*, and *I* compartments have been stratified into two subgroups to capture the dynamics of the social distancing protocols recommended by public health officials. Specifically, individuals that are not abiding by social distancing regulations, referred to in this work as “mixers”, are denoted by subscript *M*. Individuals who are following social distancing guidelines, referred to in this work as “distancers”, are denoted by subscript *D*. We assume, at the outset, that a constant proportion of individuals, denoted by *θ*, practice social distancing while social mixers make up a proportion (1 − *θ*) of the population.

The set of coupled ODEs describing the proposed Distancing-SEIRD model are given by:

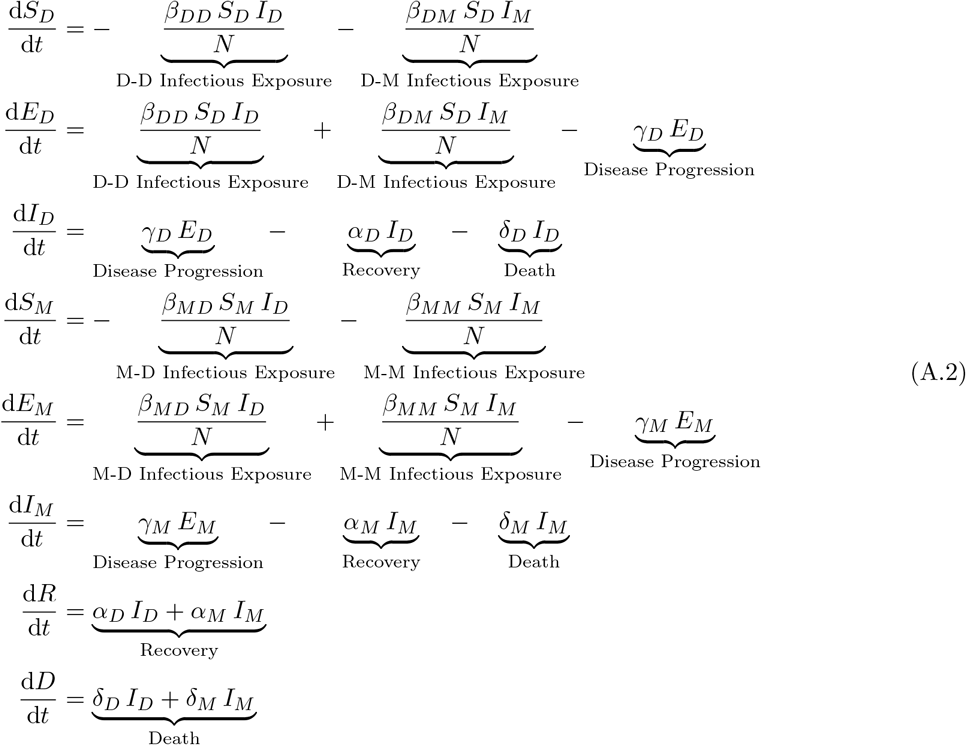

In the proposed model, the parameters *β*_*DD*_, *β*_*DM*_, *β*_*MD*_, and *β*_*MM*_ have been introduced to represent the rate of virus transmission from infected to susceptible individuals, while taking into account the social distancing practices. Specifically, *β*_*DD*_, controls the rate of transmission from infected distancers to susceptible distancers, *β*_*DM*_ from infected distancers to susceptible mixers, *β*_*MD*_ from infected mixers to susceptible distancers, and *β*_*MM*_ from infected mixers to susceptible mixers. Intuitively, individuals who practice social distancing are less likely to come into contact with others and transmit the virus. Thus we assume that *β*_*DD*_ *< β*_*DM*_ = *β*_*MD*_ *< β*_*MM*_. These assumptions imply that individuals who practice social distancing are least likely to pass the disease to one another and that individuals who do not practice social distancing are most likely to pass the disease to one another. Furthermore, by assigning *β*_*DM*_ = *β*_*MD*_, we are making the simplifying assumption that a susceptible distancer is equally likely to come into contact with an infectious mixer as a susceptible mixer is to come into contact with an infectious distancer. Moreover, we assume that this cross-compartment transmission rate falls between the two inter-compartment transmission rates. Similarly, while the pairs of parameters *γ*_*M*_ and *γ*_*D*_, *α*_*M*_ and *α*_*D*_, and *δ*_*M*_ and *δ*_*D*_ are theoretically different, we assume that each pair is the same across compartments. That is, we assume that *γ*_*M*_ = *γ*_*D*_, *α*_*M*_ = *α*_*D*_, and *δ*_*M*_ = *δ*_*D*_ and drop the subscripts. Effectively, we are assuming that the behaviour of social distancing has minimal effect on the values of these parameters. That is to say, we assume that the latent period *γ*^−1^ and the mean infectious length (*α* + *δ*)^−1^ are functions entirely of the disease and are not influenced by behaviour.

We note that the Distancing-SEIRD model developed in this work is a first generation model. Consequently a simplistic approach is implemented to model social distancing and disease transmission, and this simplistic approach is not completely accurate. The models used in this work do not included age, population density and distribution, environmental conditions, or asymptomatic individuals. Furthermore, both models consider only a single strain of the virus. Recent epidemiological data indicates that the ancestral strain of the virus comprises approximately 30% of cases, while a mutated form accounts for the remaining 70% of infections. Moreover, for simplicity, we have made the assumption that the proportion of individuals who are social distancing is a constant. In a future work, we will construct a more comprehensive model that addresses these limitations to improve the prediction accuracy of the model.

In contrast to other models that have been proposed to model the effect of social distancing and self isolation (see, for instance, [11]), we consider two infection pathways with (weak) interaction between one another. That is to say, the distancing compartment does not represent a strict quarantine in this work as we assume that social distancing individuals are still engaging in public life, just in a reduced capacity.

## B Data and Parameter Estimation

### B.1 Data Sources

The data set we use is via the repository collected by Johns Hopkins CSSE [8]. The data reported includes daily counts of confirmed cases, deaths, and daily counts of people who have recovered from COVID-19. This data used in this work was recorded from January 22, 2020 to April 13, 2020. The data set includes data for 184 countries and, for some countries (such as Canada), it also includes data at the provincial/state level. For the Canadian provincial data, the data set does not present the recovered cases individually for each province. However, they do present these counts at the national level.

For a Canadian province we estimate the recovered cases by first calculating the national time dependent case fatality rate *F*_*t*_

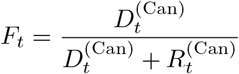

and using this number to estimate the current cumulative recovered cases via the current cumulative fatalities by

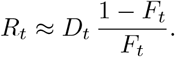

Note that for a trajectory of the SEIRD Model, *I*(*t*) represents the number of infected people at time *t* while from our data time series *I*_*t*_ represents the total cumulative number of confirmed infectious cases up to time *t*.

### B.2 Parameter Estimation

The SEIRD Model has a disease-free equilibrium at (*S, E, I, R, D*) = (*N*, 0, 0, 0, 0). If we linearize around this equilibrium, the state equations for *E* and *I* become

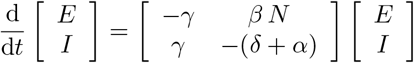

where the Jacobian matrix has leading eigenvalue

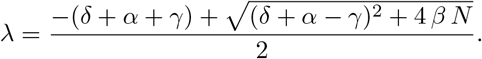

Hence *E*(*t*) and *I*(*t*) behave asymptotically exponentially around the disease free equilibrium with exponential rate given by *λ*. Thus if *λ >* 0 an outbreak occurs and if *λ <* 0 an outbreak does not occur. Since epidemic infections initially grow exponentially this *λ* can be estimated from the slope of the log-plot of infectious individuals early in an epidemic outbreak. Hence, if we can independently estimate *γ, δ*, and *α* we can use this estimate of *λ* to acquire an estimate of *β* according to the formula

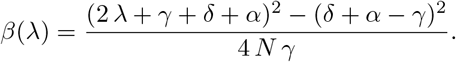

The case fatality rate represents the proportion of individuals who died from the infection relative to the the total number of individuals who had the infection. Using the notation that *D*(∞) = lim_*t*→∞_ *D*(*t*) and *R*(∞) = lim_*t*→∞_ *R*(*t*), then the case fatality rate can be represented by

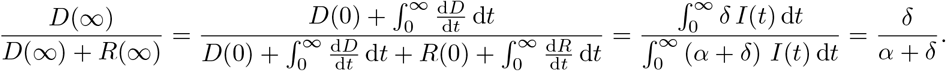

We estimate the case fatality rate for Ontario and Quebec by consider the mean value of the time dependent case fatality rate *F*_*t*_ taken over the final week of the data (April 6, 2020 to April 13, 2020 inclusive). Hence, given the case fatality rate *φ*, we can relate parameters *δ* and *α* via

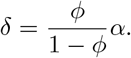

As a result, model parameters *β* and *δ* are determined from the time series data directly. The remaining model parameters, *α* and *γ*, as well as the initial number of exposed and infected individuals, *E*(0) and *I*(0), are found via an optimization procedure.

Recall that the time series *I*_*t*_ represents cumulative infections, we can estimate the current infections at time point *t* by *I*_*t*_ − *R*_*t*_ − *D*_*t*_. Similarly, for a trajectory of the SEIRD Model we can represent the cumulative infections at time *t* by *I*(*t*) + *R*(*t*) + *D*(*t*). In fitting the data, in practice, we utilize both estimates.

Let

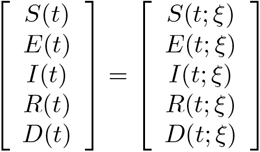

represent a trajectory of the SEIRD Model A.1 parameterized by *ξ* = (*E*_0_, *I*_0_, *γ, α*) with initial condition

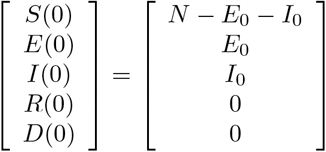

where the population size *N* is treated as a known, fixed value. The optimization problem is to find 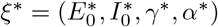 that minimizes the cost function *χ* given by

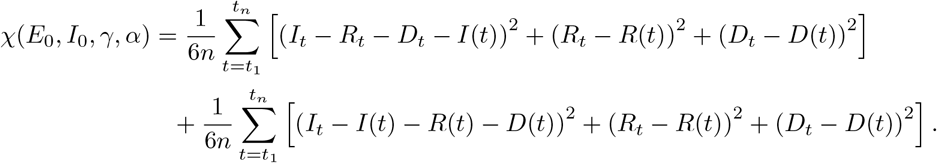

Note that *χ* then represents the mean squared errors of fitting (*I*(*t*), *R*(*t*), *D*(*t*)) to (*I*_*t*_ −*R*_*t*_ −*D*_*t*_, *R*_*t*_, *D*_*t*_) and fitting (*I*(*t*) + *R*(*t*) + *D*(*t*), *R*(*t*), *D*(*t*)) to (*I*_*t*_, *R*_*t*_, *D*_*t*_).

We provide reasonable bounds for this fitting process by assuming that 0 *≤ E*_0_, *I*_0_ *≤* 20, 10^−1^ ≤ *γ ≤* 0.8, and 20^−1^ *≤ α ≤* 0.3. In practice, the bounds we set for *E*_0_ and *I*_0_ are likely too large, however restricting the bounds did not change the optimizer discovered. The *γ* bound was chosen based on reports that the latent period of the disease has a median value of 5.1 days with a 95% Confidence Interval of 4.5 to 5.8 days [12]. We chose our range for *γ*^−1^ to be between 1.25 days and 10 days, in order to ensure that we capture the most likely parameter values. Similarly, the bounds on *α* were chosen based on reports that the mean infectious period is around 7-14 days (for instance, in [13] they suggest a quarantine time of 14 days from first symptom onset, in order to prevent further infections). Like with *γ*, we chose the bounds here to be larger than necessary in order to capture the most likely parameter value. Other reports provide ranges for the latent period and mean infectious period that fall within these bounds, see [14, 15, 16, 17].

With these ranges set, we approximate the optimizer via Gaussian maximum likelihood estimation with differential evolution minimization [18]. The final result is then further optimized within our provided bounds by using the L-BFGS-B method from SciPy [19]. The quality improvements of the fit after this final local optimization step are minor, indicating that differential evolution of the maximum likelihood estimator already achieves accurate parameter discovery. We numerically solve the differential equation system with the `integrate.odeint` function from SciPy that acts as a wrapper for `lsoda` [19, 20].

The results of this process for China, Canada, Ontario, and Quebec are provided in Table 1. Note, when fitting the China data we widened the bounds of *E*_0_ and *I*_0_. Since the reported data begins on January 22, 2020 at which point there were already 548 infections, we extended the bounds in this case to be 0 *≤ E*_0_, *I*_0_ *≤* 1000.

### C Relation between transmission rates in model 1 and model 2

Fitting the SEIRD Model (Eq. (A.1)) to data, as described in Section B.2, results in a single *β* value. Now we discuss how this single, effective *β* value relates to the three parameters *β*_*DD*_, *β*_*DM*_, and *β*_*MM*_ from the Distancing-SEIRD Model (Eq. (A.2)). In order to compare the exposed and infected compartments between the two models, define *Ê* = *E*_*D*_ + *E*_*M*_ and *Î* = *I*_*D*_ + *I*_*M*_. Hence,

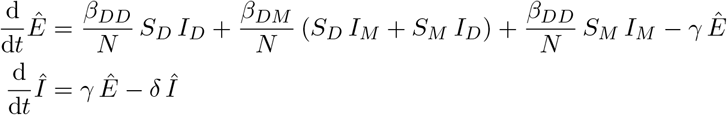

The Distancing-SEIRD Model has a disease-free equilibrium corresponding to *S*_*D*_ = *θ N, S*_*M*_ = (1 −*θ*) *N* and *I*_*D*_ = *I*_*M*_ = *E*_*D*_ = *E*_*M*_ = *R* = 0. Consider linearizing Eq. (A.2) about this disease-free equilibrium. In the neighbourhood of this disease-free equilibrium note that *S*_*D*_ and *S*_*M*_ are effectively constant (up to a super-linear correction), hence, taking *S*_*D*_ = *θ N* and *S*_*M*_ = (1 − *θ*) *N*

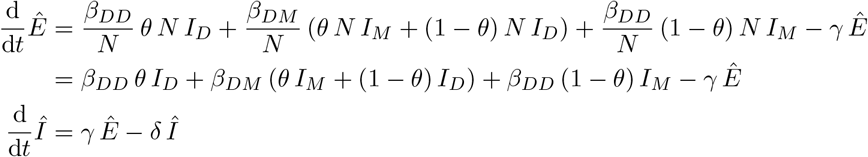

also note that in the neighbourhood of this equilibrium, *I*_*D*_ = *θ Î* and *I*_*M*_ = (1 *θ*) *Î* (up to a super-linear correction), hence

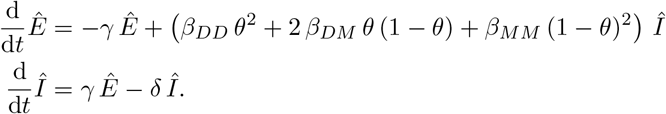

Now comparing this with the linearization of the SEIRD Model in Section B.2, we find that the two systems are asymptotically equivalent under the change of variables

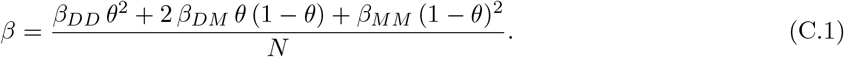

Hence, when modelling a particular outbreak where the value of *β* for an SEIRD model has been determined, one can use Eq. C.1 to determine appropriate values of *β*_*DD*_, *β*_*DM*_, and *β*_*MM*_ such that their combined effect still behaves appropriately. For the numerical experiments discussed in Section 3, for a given social distancing proportion *θ* and a given data informed *β*, we determined the values of *β*_*DD*_, *β*_*DM*_, and *β*_*MM*_ by using Eq. C.1 along with the heuristic assumption that *β*_*DD*_ = *β*_*MM*_ */*100 and *β*_*DM*_ = *β*_*MM*_ */*20. The assumption on *β*_*DD*_ is equivalent to assuming that distancing individuals are only 1% as likely to transmit the virus to one another as mixing individuals are. Likewise the assumption on *β*_*DM*_ is that the distancing and mixing individuals are only 5% as likely to transmit the virus to one another as mixing individuals are. These effects are simplifying assumptions and other ratios could have been chosen. An important piece to note is that *β*_*DD*_ = 0. We do not consider social distancing to be the same as complete isolation. This is meant to capture the fact that social distancers will still interact with the general public either by visiting the grocery store or the doctor or in some other reasonable exposure. With these ratios between the compartmental transmission rates, we can determine the final value of *β*_*MM*_ (and thus *β*_*DD*_ and *β*_*DM*_ as well) for a given *θ* by solving Eq. C.1.

